# Data Quality of Out-Of-Pocket Payment on Institutional Delivery in India

**DOI:** 10.1101/2023.04.18.23288434

**Authors:** Sanjay K. Mohanty, Laxmi Kant Dwivedi, Santosh Kumar Sharma, Sowmya Ramesh, Priyanka Gautam, Suraj Maiti, Saritha Nair, S. K. Singh

## Abstract

Estimates of out-of-pocket (OOP) payment on health care are increasingly used in research and policy. In India, estimates of OOP payment are usually derived from health surveys carried out by the National Sample Survey (NSS). The questions on OOP payment on delivery care have recently integrated in the last two rounds of India’s National Family and Health Survey (NFHS-4 & NFHS-5). There are several issues relating to design of questions, reporting and recording of responses that have bearing on reliability of OOP estimates. This paper compares the OOP estimates from latest rounds of two of the large-scale population-based surveys; NFHS-5, 2019-21 and the National Sample Survey (NSS), 2018. We also highlight the type of question canvassed and its implications on OOP estimates of NFHS-5 survey. We used 155,624 births that were reported between in NFHS-5 and a total of 27,664 hospitalised cases for delivery care that were recorded in 75^th^ round of NSS health survey, 2018. We have used descriptive statistics and two-part regression model to examine variations of OOP across surveys. We found large variations in distribution of OOP payment in NFHS-5 and NSS survey. Based on births during the five years preceding the survey, the OOP payment on institutional birth from public health centres in India from NFHS-5 was INR 2,894 (95% CI:2843-2945) compared to INR 2,738 (95% CI: 2644-2832) from NSS. Variations are similar for those availing services from private health centres. Controlling for socio-economic and demographic characteristics, the OOP payment from NFHS was lower among poorest and higher among richest compared to NSS. The variations in OOP across two surveys were larger across states of India. The variations in OOP payment across surveys were possibly due to structure of questions, recall bias, and variations in price level. We suggest to canvass standardised questions across surveys to obtain reliable OOP estimates across surveys.

## Introduction

Indicators of financial protection are increasingly sought in global, national and local developmental programme. Many national and regional government and developmental partner are now collecting these indicators through population-based surveys. Reliability of these estimates is paramount for guiding research and policy. Measurement error in population-based surveys that may arise due to the wording, length and reference period affect reliability of estimates. Interviewer and respondent bias such as incorrect reading/understanding of questionnaire, wrong recording of responses, missing data and memory loss of respondent also affect data quality (Visaria 1980; Bradburn et al. 1987; Scott 1990; Biemer 1991; Branch 1994; Anand and Harris 1994; Bradburn 2004). Estimates may be under-reported or over reported due to one or more of these factors.

India’s National Health Mission (NHM)–the largest publicly-funded health care program worldwide – has recognized the OOP payment as a major barrier to institutional delivery in the country. It implements conditional cash transfer schemes, such as the *Janani Suraksha Yojana (JSY) and the Janani Shishu Suraksha Karyakram (JSSK)*, to reduce high OOP payment and CHE on maternal care (MoHFW; 2005). In last 15 years, the NHM has doubled the coverage of institutional delivery (from 38.7% in 2005-06 to 89% by 2019-21), reduced inequality in utilization of institutional delivery, and reduced infant, neonatal and maternal mortality in the country (Ali et al., 2020; Ghosh, 2019). Recently, the India’s Demographic and Health Survey, the National Family and Health Survey (NFHS 4, NFHS 5) has integrated questions on OOP payment on delivery care and generate evidences for research, programme and policy.

A decade ago, the NSS was the only source that collected nationwide data on medical expenditure and institutional delivery through its household health survey and estimates were provided across the states and union territories of India (NSSO, 2006, 2015, 2018). Expenditure and reimbursement on maternal care, used to estimate OOP has advantages of being numerically measured. Disaggregated data on expenditure of delivery such as expenditure on hospital stay, tests, medicines, transportation costs, and other costs and reimbursement were collected as a part of hospitalization. Respondents who could not report expenditure on the individual components were asked to provide the total expenditure on delivery care. Estimates of OOP are derived based on expenditure and reimbursement. The NSS data has been extensively used among researcher for estimation of OOP and CHE and in national and state policy (Goli, 2016; Joe, 2015; Kastor & Mohanty, 2018; Mohanty et al., 2020; Pandey et al., 2018a; Pandey et al., 2018; MoHFW 2017). A few studies based on District Level Household Survey (DLHS) data have highlighted the variations in institutional delivery in India (Modugu, 2012; Mohanty & Srivastava, 2013; Roy, 2018; Vellakkal, 2017).

The fourth and fifth rounds of the National Family Health Survey (NFHS) collected data on OOP payment on institutional delivery from mothers who had recently delivered. An increasing number of studies have been using this data to estimate maternal expenditure (Kodali, 2019; Mohanty et al., 2020; Mohanty et al., 2019; Rout, 2019). Questions of OOP payment were asked direct which may not have been easy for the respondents to report. Besides, data on OOP were based on the last births to mothers during five years preceding the survey.

Though, both NFHS and the NSS surveys yield estimates of OOP payment on maternal care, but they differ in structure and design of questions. While data on these indicators are now available and used at states and district level, little is known on comparability and reliability of the estimates in India. Given the demand for these evidences for research and policy, this paper examines the issues pertaining to data quality of OOP payment of institutional delivery from NFHS-5, 2019-21 by comparing with the estimates from the 75^th^ round of NSS, 2018. It also suggests possible way to avoid the issues and provide reliable estimates of OOP in India.

## Materials and Methods

### Materials

We used the unit data from two large-scale population-based surveys, namely, the NFHS-5, conducted during 2019-21, and the 75^th^ round of the health survey (25.0 schedule), 2018 carried out by the National Sample Survey. These are secondary datasets which are available in the public domain for research purposes. Both the surveys were population representative and provided data on our variable of interest: out-of-pocket payments on institutional delivery. A brief description of the data structure of both the surveys is given below.

The NFHS-5 collected compressive information on maternal and child health across the states and union territories of India. In the survey a stratified two stage sampling design was adopted to provide reliable estimates at national, state and districts of India. In rural areas, census villages and in urban areas, the census enumeration block (CEB) are used as the primary sampling unit (PSU). The urban and rural areas were selected using Probability Proportional to Size (PPS) systematic sampling. In total, 30,198 PSUs were surveyed across the country. The survey successfully interviewed 636,699 households and 552,040 ever-married women, in the age group 15-49, and 101,839 men, in the age group 15-54, across the states and union territories of India. Details on the sampling design, coverage, and findings of the survey are available in the national report (IIPS & ICF 2021). We have used the unit data from the kids file, which provides details of births to mothers during the five years preceding the survey. Of the 232,920 births during that period, 1,76,843 were the last births to mothers, of which, 155,624 took place in health facilities. NFHS-5 collected data on OOP payment on the last births during a period of five years. For the first time, NFHS-4 included a new set of questions on OOP payment on institutional delivery and continued in NFHS-5. The data on OOP payment was collected through a set of six questions that pertained to OOP payment on delivery by hospital stay, tests, medicines, transportation, and other costs in five year preceding the survey. Accordingly, the mid-year of estimates of NFHS-5 would be close to 2018, the same year NSS data refers to. Respondents were asked, “*How much was the out of pocket cost for transportation/ hospital stay/ medicines, etc*.*?*” (Question no. 448a, 448ba, 448bb, 448bc, 448bd, 449) (S1(a) Table). Respondents who could not report OOP payment on the individual components of delivery were asked to describe the total OOP payment on delivery care.

The 25^th^ schedule of the 75^th^ round of the health survey, 2018 (henceforth referred to as 75(25.0)), covered a sample of 113,823 households and 555,352 individuals. The survey adopted a stratified, multistage cluster sampling design. The first stage units (FSU) are the census villages in the rural areas and urban frame survey (UFS) in urban areas. The sample villages in both urban and rural areas have been selected based on Probability Proportional to size With Replacement (PPSWR). A total of 14, 300 FSU were covered at all India level. The sampling methodology and the findings of the survey are available in the respective reports (NSSO, 2018). The primary focus of the NSS-based health survey was to provide comprehensive information on health expenditure, hospitalization, outpatient visits, type of ailment, nature of ailment, total duration of ailment, expenses on pre- and post-natal care, and so on. The question on health expenditure was systematically canvassed for each episode of hospitalization/outpatient visit, along with type of ailment, level of care, means of meeting health care expenditure, etc. Information on delivery care expenditure was collected as a part of hospitalization within a reference period of 365 days prior to the surveys (S1(b) Table) The information was collected at a disaggregated level across eight sub-components: package components, doctor’s or surgeon’s fee (hospital staff or other specialists), medicines, diagnostic tests, bed charges, other medical expenses (attendant charges, charges related to physiotherapy, personal medical appliances, blood, oxygen), transportation costs for patient, other non-medical expenses incurred by household on food, transportation costs for others, expenditure on escort, and lodging charges if any. The OOP payment was derived from the total expenditure and reimbursement and estimated in a reference period of 365 days. Of the 91,449 individuals hospitalized during the 365 days prior to the survey, 27,664 women reported having a hospital birth.

## Variables

### Outcome variable

Out-of-pocket payment on institutional delivery and its components (hospital stay, tests, medicines, transportation, others) was the dependent variable used in the analyses.

### Independent variables

A set of independent variables were used in the analysis. These include place of delivery (public, private), place of residence (rural, urban), maternal education (illiterate, primary, middle/secondary, higher secondary & above), household size (1-4, 5-7, 8+), wealth/MPCE quintile (poorest, poorer, middle, richer, richest), religion (Hindu, Muslim, Others), (scheduled caste/scheduled tribe (SC/ST), other backward classes (OBC), Other), states, etc.

### Methods

Descriptive statistics was used to quantify the changes in the outcome variable. Estimates of mean are supported with 95% confidence intervals. The distribution of OOP payment from NFHS and NSS surveys across economic conditions of household and type of health centers were presented with help of density plots.

### Test for Equivalence

The objective of our analyses is to show the comparability between the estimates from NFHS and NSS surveys, both of which are population representative. We have employed the Two One Sided Test (TOST) Procedure (Schurimann, 1987; Anderson and Hauck, 1983) that used as a test of non-equivalence. We have employed the test in Stata by using the user written “tostti” package. The main rationale in using TOST procedure is that it is a test that “fits for all” in a general sense.

### Multivariate analysis

We have used the two-part regression model to predict the OOP payment of institutional delivery after adjusting for selected socio-demographic and economic variables for both NFHS-5 and NSS. The OOP payment (dependent variable) contains a large number of zeroes which is also a generic feature of health expenditure and cost data worldwide. Existing literature suggest two-part model as appropriate statistical tool and best suited in such cases because of followings (Deb, P et.al 2018; Garcia et.al 2013; Humphrey, 2013). Firstly, the variable of concern, OOP has genuine zeroes under its aegis. Second, if the zeroes are true ones and also there is sequential ordering of events in the outcome variable, like in this case, incurrence of OOP happens after institutional delivery, it is better to use two-part model. Thirdly, there might be different models available to address skewness and zeroes, but there is no unique model that is capable of dealing with all issues simultaneously (Gregori et al. 2011; Mihaylova et al. 2011). The choice of model depends on the data that we are using (Gregori et al. 2011), and simple models are much more effective than their complex counterpart (Mihaylova et al. 2011). Taking into consideration the facts like true nature of the zeroes, sequential occurrence of the outcome variable and ease of interpretability, we used “Two-Part Models”.

The analysis was done using “twopm” package in Stata version 16.0. The first part of our analysis, using a logistic model, described the likelihood of an individual incurring OOP payment on institutional delivery by selected socio-demographic and economic variables.

The model took the following formxs

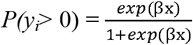

where *y*_*i*_= 0 indicates that there was no OOP payment on institutional delivery.

The second part of the model uses ordinary least square (OLS) regression to determine the probability of incurring any OOP payment on institutional delivery. The OLS regression model used the logarithm of OOP payment on institutional delivery as a dependent variable. The model predicted the OOP payment on institutional delivery after adjusting for selected socio-demographic and economic variables.

## Results

Table 1 presents the characteristics of sample respondents from NFHS-5 and NSS surveys. Both NFHS-5 and NSS are large-scale population-based representative surveys and provide expenditure on institutional delivery. The key indicators, such as median age, average household size, proportion of urban population, Hindus, and scheduled caste and scheduled tribe population, were used to show how similar both the surveys are, as the cse should be because the two surveys are based on the same population. The proportion of institutional deliveries in public and private health centres was also similar in both the surveys [68.5% & 31.8% respectively in NFHS-5 and 70.2% & 29.8% respectively in NSS 75^th^ round]. The average expenditure on institutional birth at current prices was INR 9,294 in NFHS-5 compared to INR 8,904 in 2018. We can say that the estimates are not equivalent or similar, because there exist relevant differences as observed from the TOST test.

**Table 1:** Sample characteristics of respondents surveyed in NFHS-5, 2019-21 and NSS, 2018.

|  | NFHS-5, 2019-21 | NSS 75th round, 2018 |
| --- | --- | --- |
| Number of institutional births | 155,624 | 27,664 |
| Median age of mothers (year) | 26 | 26 |
| Average household size | 6 | 6.0 |
| % Urban | 29.65 | 25.65 |
| %SC/ ST | 31.54 | 30.50 |
| %Hindus | 80.27 | 79.28 |
| Share of institutional delivery at public health centre (%) | 68.53 | 70.24 |
| Share of institutional delivery at private health centre (%) | 31.47 | 29.76 |
| OOP payment on institutional birth [95%CI] (INR) | 9294[9150-9438] | 8904[8421-9387] |
| OOP payment on birth at public health centre[95%CI] (INR) | 2894[2843-2945] | 2738[2644-2832] |
| OOP payment on birth at Private health centre[95%CI] (INR) | 23,231[22906-23557] | 23,460[22141-24779] |

In Figure 1, we have plotted the distribution of log transformed OOP payment for NFHS-5, 2019-21 and of NSS, 2018. It is observed that the distribution of OOP payment for NFHS is more skewed and kurtic than that of NSS. Heaping and digit preference are relatively higher in NFHS compared to NSS. There exists prominent crests and troughs in the plot of OOP payment for NFHS whereas, in case of NSS, the histogram is much smoother and resembles the Normal density.

**Figure 1.** Density plot of log transformed OOP payment, NFHS-5 and NSS, 2018.

S1 Fig. and S2 Fig. shows the weighted log transformation of OOP on institutional delivery based on NFHS-5, 2019-21 and NSS health survey, 2018. These distributions are individually compared with Normal Density curve. We observed that the distribution of OOP payment on institutional delivery is smoother and close to normality for NSS as compared with NFHS.

In Figure 2, we have plotted the distribution of log transformed OOP payments for NSS, 2018 and NFHS, 2019-21 by place of delivery at public and private health centers. Those delivered at private health centers, the OOP payment was much higher for NFHS compared to that of NSS. On the contrary, for those delivering at public health centers, we observe the reverse. Most of the observations were distributed towards the left of the NSS distribution suggesting that NFHS here is underestimating the OOP payments as compared to NSS.

**Figure 2:** Density plot of log transformed OOP payment by place of institutional delivery, NFHS-5 and NSS, 2018.

In Figure 3, we have plotted the density of log transformed OOP payment for NSS, 2018 and NFHS, 2019-21 by economic conditions of households, viz., poorest, poorer, middle, richer and richest. In general, there is differences in the skewness and kurtosis of the distribution of OOP payment for NFHS compared to NSS in poorest and richest quintile. For the poorer, middle, and richer, there is some deal of overlapping in the distribution from these two surveys. In case of poorest, the NFHS density plot of OOP tend towards the lower end and lies below NSS in most part. In case of the richest category, we observe that NFHS overestimated the OOP payment for higher levels of income within the category.

**Figure 3:** Density plot of log transformed OOP payment based on wealth, NFHS-5 and MPCE quintile, NSS, 2018.

Table 2 presents the estimates of OOP payments for institutional delivery from NFHS-5 and NSS survey. In NFHS surveys, we find a systematic pattern in deviation of OOP payment from NSS by economic conditions of households. In general, the OOP payment from NFHS for the poorest and poorer were significantly lower while that of richer and richest are significantly higher than that of NSS estimates. For instance, in the poorest uintile, the NFHS estimates were lower by 28%, 12% for poorer, higher by 6% in case of richer and 22% higher in case of richest. Based on these deviations, we can say that the NFHS estimates at lower economic strata were underestimates and for higher economic strata were overestimates (NSS as a gold standard). The pattern was also similar for educational attainment. The estimates of OOP payment on institutional delivery by education varied from INR 4,472 [95% CI: 4315-4630] among women with no education to INR 17,805 [17369-18241] among women with higher education in NFHS-5. Estimates based on NSS 75^th^ round varied from INR 4,716 [4115-5317] among women with no education to INR 16,479 [14956-18001] among women with higher education. Differences of OOP payment from NFHS and NSS was 5% among women with no education and 8% for those with higher secondary and above. The TOST test has been done for each of the estimate of mean of OOP payments across the SES variables. Though the 95% confidence intervals are overlapping in many of the mean estimates, we conclude from the test of equivalence that, for most of the mean estimates there exists relevant differences, at levels of Rs. 250, Rs. 500 and Rs. 750. This implies, that in the two surveys there are significant difference in mean OOP

**Table 2:**
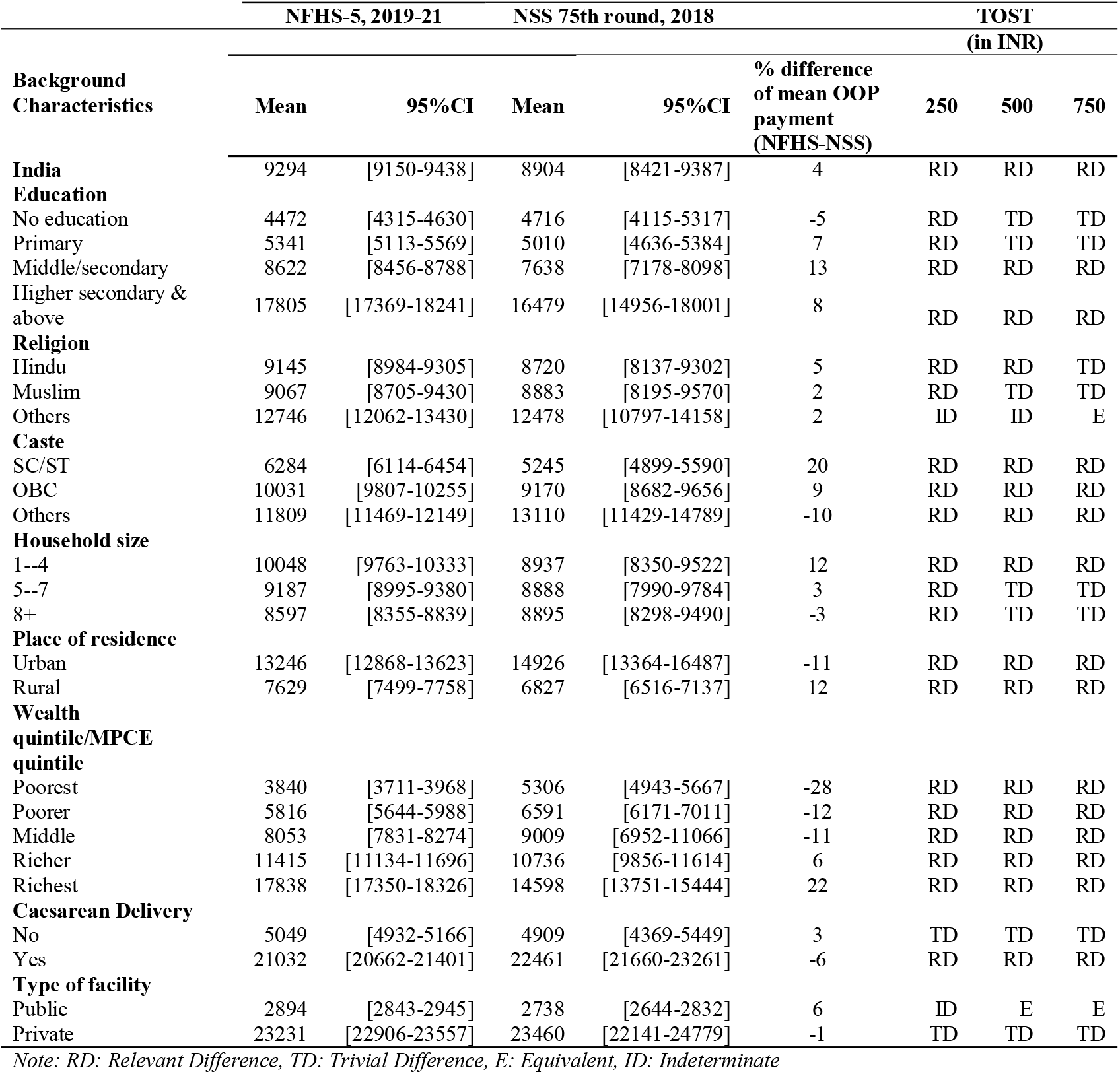
Mean OOP payment on institutional delivery background characteristics in India.

Figure 4 presents the inter-state variations in the estimates of OOP payment of institutional delivery. Several states showed lower estimates of OOP payment for institutional delivery in NFHS-5 compared to NSS survey. Estimates were lower in the states of Lakshadweep, Haryana, Chandigarh, Telangana, West-Bengal, Puducherry, Delhi, Andhra Pradesh, Rajasthan, Kerala, Karnataka, and the north-eastern states. Similarly, several states showed higher OOP in NFHS-5 compared to NSS survey. Table 3 presents the point estimates of OOP payments and 95% confidence interval of institutional delivery from NFHS-5 and NSS surveys. The variations in the OOP payment on institutional delivery from NFHS-5 and NSS, 2018 health surveys were large across states of India. The mean OOP payment on institutional delivery from NFHS-5 were lower than NSS estimates in 14 states and higher in 21 states. The difference in the estimates varied from 129% in Arunachal Pradesh, 85% in Manipur, 82% in Mizoram, and 62% in Meghalaya. It was lower by 38% in Lakshadweep, 36% in Haryana, 27% in Chandigarh and 17% in Telangana. The 95% confidence interval are non-overlapping in the states of Chhattisgarh, Jharkhand, Assam, Bihar, Madhya Pradesh, Meghalaya, Mizoram, Manipur and Arunachal Pradesh.

**Figure 4:** Out-of-pocket expenditure (INR) on institutional delivery by major states from NFHS-5 and NSS, 2018.

**Table 3:** Mean OOP payment on institutional delivery (INR) by major states of India.

| States | NFHS-5 (2019-21) |  | NSS 75th round, 2018 |  | % difference<br>of mean<br>OOP<br>payment |
| --- | --- | --- | --- | --- | --- |
|  | Mean | 95%CI | Mean | 95%CI |  |
| **India** | 9294 | [9150-9438] | 8904 | [8421-9387] | 4 |
| Daman & Diu | 5810* | [4322-7298] * | 10609 | [2442-18776] |  |
| Dadra & Nagar Haveli |  |  | 2182 | [637-3728] | -45 |
| Lakshadweep | 13005 | [8088-17921] | 20862 | [10633-31091] | -38 |
| Haryana | 10095 | [9512-10679] | 15802 | [2332-29271] | -36 |
| Chandigarh | 9442 | [5432-13452] | 12920 | [7063-18776] | -27 |
| Telangana | 14009 | [13181-14837] | 16913 | [14480-19346] | -17 |
| West Bengal | 6671 | [6215-7126] | 7903 | [7049-8757] | -16 |
| Pondicherry | 8291 | [6519-10064] | 9643 | [5745-13541] | -14 |
| Delhi | 12306 | [11086-13525] | 14230 | [10395-18065] | -14 |
| Tripura | 7454 | [6673-8234] | 8333 | [6871-9794] | -11 |
| Andhra Pradesh | 12038 | [11183-12893] | 13415 | [11923-14907] | -10 |
| Rajasthan | 4878 | [4616-5140] | 5282 | [4712-5853] | -8 |
| Kerala | 22651 | [21375-23926] | 23861 | [22293-25429] | -5 |
| Karnataka | 12316 | [11570-13061] | 12628 | [11403-13853] | -2 |
| Jammu & Kashmir | 6732 | [6313-7151] | 6782 | [6034-7531] | -1 |
| Tamil Nadu | 14856 | [14032-15679] | 14760 | [13104-16416] | 1 |
| Maharashtra | 12114 | [11264-12963] | 11843 | [10752-12934] | 2 |
| Uttar Pradesh | 7981 | [7722-8241] | 7506 | [6564-8448] | 6 |
| Sikkim | 8956 | [7322-10590] | 8182 | [6472-9892] | 9 |
| Himachal Pradesh | 8654 | [7269-10039] | 7690 | [6225-9156] | 13 |
| Gujarat | 10147 | [9481-10812] | 8919 | [7836-10001] | 14 |
| Punjab | 13895 | [13072-14718] | 12136 | [10923-13350] | 14 |
| Uttarakhand | 12391 | [11164-13618] | 10467 | [8407-12527] | 18 |
| Andaman & Nicobar Islands | 8391 | [5317-11465] | 7029 | [3770-10287] | 19 |
| Nagaland | 8310 | [7309-9312] | 6954 | [4720-9188] | 19 |
| Orissa | 7523 | [7111-7935] | 6234 | [5567-6901] | 21 |
| Chhattisgarh | 6030 | [5494-6567] | 4579 | [3723-5436] | 32 |
| Jharkhand | 7054 | [6444-7665] | 5285 | [4391-6178] | 33 |
| Assam | 8397 | [7952-8843] | 6264 | [5251-7276] | 34 |
| Bihar | 6973 | [6605-7342] | 5179 | [4556-5802] | 35 |
| Goa | 18370 | [15099-21641] | 12965 | [8713-17217] | 42 |
| Madhya Pradesh | 5017 | [4675-5359] | 3494 | [2934-4054] | 44 |
| Meghalaya | 5741 | [4725-6757] | 3535 | [2850-4220] | 62 |
| Mizoram | 4936 | [3958-5914] | 2717 | [2259-3176] | 82 |
| Manipur | 20953 | [19775-22131] | 11304 | [10200-12409] | 85 |
| Arunachal Pradesh | 10709 | [9607-11811] | 4684 | [3625-5743] | 129 |
| Ladakh | 4089 | [3568-4611] |  |  |  |
| **Total** | **1,55,624** |  | **27,664** |  |  |

\* Daman & Diu and Dadar & Nagar haveli are combined in NFHS-5 raw data, hence the estimates are combined

Table 4 presents the percentage share of the components of OOP payment on institutional delivery from NFHS-5 and NSS 75^th^ round of surveys according to some selected background characteristics in India. In India, the share of expenditure on hospital stay, diagnostic/tests, medicines and other costs in the overall OOP payment was 31.2%, 16.4%, 20.5% and 31.9% respectively in NFHS-5, whereas it was 30.2%, 17.9%, 32.2%, 19.7% respectively in NSS 75^th^ round, which can be seen in Figures 5 and 6 respectively. The share of OOP payment for hospital stays had strong education gradient in both the surveys; however, the percentage was higher in NFHS-5 compared to NSS 75^th^ Round. The share of OOP payment on hospital stays (29.7% vs. 27%) and other costs (33.5% vs. 21.7%) in urban areas was lower in NSS as compared to NFHS-5. The share of OOP payment on hospital stays, tests, and medicines across wealth quintiles was higher in NSS than in NFHS-5. In case of hospital stays, the OOP payment on institutional delivery in private health centres was higher than in public health centres in both the surveys. In NFHS-5, about 36% of OOP payment went towards hospital stay in private health centres, whereas the corresponding figure was about 41.2%in NSS 75^th^ round.

**Table 4:** Percentage share of components of OOP payment on institutional delivery by background characteristics.

| Background Characteristics | NFHS-5, 2019-21 |  |  |  | NSS 75th round, 2018 |  |  |  |
| --- | --- | --- | --- | --- | --- | --- | --- | --- |
|  | Hospital stay (%) | Test Done (%) | Medicine (%) | Other cost (%) | Hospital stay (%) | Test Done (%) | Medicine (%) | Other cost (%) |
| **Education** |  |  |  |  |  |  |  |  |
| No education | 23.7 | 13.8 | 22.2 | 40.2 | 23.8 | 18.4 | 36.2 | 21.5 |
| Primary | 26.4 | 14.3 | 21.6 | 37.7 | 22.1 | 19.0 | 36.1 | 22.8 |
| Middle/secondary | 30.8 | 16.2 | 21.0 | 32.1 | 28.1 | 17.2 | 33.4 | 21.3 |
| Higher secondary & above | 34.3 | 17.7 | 19.5 | 28.6 | 35.8 | 17.0 | 29.5 | 17.7 |
| **Religion** |  |  |  |  |  |  |  |  |
| Hindu | 30.8 | 16.2 | 20.3 | 32.7 | 30.3 | 17.8 | 32.4 | 19.5 |
| Muslim | 32.7 | 16.2 | 21.1 | 30.0 | 29.1 | 18.4 | 32.1 | 20.4 |
| Others | 33.3 | 19.1 | 21.6 | 26.0 | 32.3 | 17.7 | 29.7 | 20.3 |
| **Caste** |  |  |  |  |  |  |  |  |
| SC/ST | 28.8 | 15.2 | 21.1 | 34.8 | 22.5 | 18.7 | 35.6 | 23.1 |
| OBC | 30.1 | 16.8 | 20.2 | 32.9 | 31.5 | 17.2 | 31.9 | 19.4 |
| Others | 34.5 | 16.6 | 20.6 | 28.3 | 33.4 | 17.9 | 30.2 | 18.5 |
| **Household size** |  |  |  |  |  |  |  |  |
| 1--4 | 30.9 | 17.0 | 20.7 | 31.5 | 31.7 | 17.5 | 31.2 | 19.6 |
| 5--7 | 31.4 | 16.2 | 20.5 | 31.9 | 29.0 | 18.8 | 32.1 | 20.2 |
| 8+ | 31.2 | 15.8 | 20.4 | 32.5 | 30.5 | 16.7 | 33.7 | 19.1 |
| **Place of residence** |  |  |  |  |  |  |  |  |
| Rural | 33.4 | 17.4 | 19.7 | 29.6 | 35.1 | 18.0 | 30.0 | 16.9 |
| Urban | 29.7 | 15.6 | 21.2 | 33.5 | 27.0 | 17.5 | 33.8 | 21.7 |
| **Wealth quintile/MPCE quintile** |  |  |  |  |  |  |  |  |
| Poorest | 23.4 | 12.4 | 24.5 | 39.6 | 24.4 | 18.5 | 35.9 | 21.2 |
| Poorer | 26.5 | 14.4 | 21.8 | 37.3 | 26.0 | 18.7 | 34.3 | 21.0 |
| Middle | 30.1 | 16.2 | 20.9 | 32.8 | 28.4 | 18.4 | 32.0 | 21.2 |
| Richer | 32.2 | 17.1 | 19.8 | 30.9 | 33.4 | 17.3 | 31.1 | 18.3 |
| Richest | 34.8 | 17.7 | 19.5 | 28.0 | 33.9 | 16.7 | 31.0 | 18.3 |
| **Type of Facility** |  |  |  |  |  |  |  |  |
| Public health facility | 14.2 | 12.9 | 25.5 | 47.4 | 5.4 | 21.1 | 40.3 | 33.2 |
| Private health facility | 36.0 | 17.7 | 19.4 | 27.0 | 41.2 | 14.3 | 28.8 | 15.7 |
| **Total** | **31.2** | **16.4** | **20.5** | **31.9** | **30.2** | **17.9** | **32.2** | **19.7** |

**Figure 5.** Percentage share of OOP payment on institutional delivery (in INR) by type in India based on NFHS-5, 2019-21.

**Figure 6.** Percentage share of OOP payment on institutional delivery (in INR) by year of birth in India based on NSS, 2018.

Table 5 presents the regression results of the two-part model and the predicted OOP payment for institutional delivery, for NFHS-5, 2019-21. The result from the logit regression revealed that the likelihood of incurring OOP payment on institutional delivery was significantly higher for those mothers with higher secondary and above education and for those belonging to other backward caste (OBC). For instance, women with higher education had 18.8% more likelihood of incurring OOP payment for institutional delivery compared to women with no education. Similarly, the likelihood of incurring OOP payment was higher among those belonging to OBC and other caste groups compared to SCs/STs. Furthermore, women who delivered in public health centres were 57% less likely (Coeff=-1.57; 95%CI=-1.64,-1.51) to incur OOP payment compared to those who delivered in private health centres. In the second part of the model, the log transformation of OOP payments, was used as the dependent variable. The probability of incurring any OOP payment on institutional delivery was higher among women with higher education, those belonging to other religious groups and castes, richest wealth quintile, and those who delivered in private health centres. The probability of incurring any OOP payment on institutional delivery was 39.1% higher among women with higher education than their other counterparts. It was found that the wealth quintile shows the negative coefficients for the logit model and positive coefficients for the OLS model. The probability of incurring any OOP payment decreased as the wealth quintile increases, whereas the amount of OOP payment increased as the wealth quintile increases. The probability of incurring any OOP payment on institutional delivery was two times higher in private health centres compared to public health centres. The year of births present negative coefficients for both logit and OLS model.

**Table 5:** Results of the two-part regression model and predicted OOPE on institutional delivery in India, NFHS-5 (2019–21)

| Background characteristics | $\beta$ (logit) | 95% CI | $\beta$ (OLS) | 95% CI | Mean OOP payment |
| --- | --- | --- | --- | --- | --- |
| **Education** |  |  |  |  |  |
| No education |  |  |  |  | 5610 |
| Primary | -0.023 | [-0.09, 0.04] | 0.048* | [0.01, 0.09] | 6392 |
| Middle/secondary | 0.03 | [-0.02, 0.08] | 0.209*** | [0.18, 0.24] | 10988 |
| Higher secondary & above | 0.188*** | [0.11, 0.27] | 0.391*** | [0.35, 0.43] | 23724 |
| **Religion** |  |  |  |  |  |
| Hindu |  |  |  |  | 11663 |
| Muslim | 0.052 | [-0.02, 0.12] | 0.021 | [-0.01, 0.05] | 12289 |
| Others | -0.150*** | [-0.24, -0.06] | 0.135*** | [0.06, 0.21] | 16554 |
| **Caste** |  |  |  |  |  |
| SC/ST |  |  |  |  | 7761 |
| OBC | 0.168*** | [0.12, 0.21] | 0.075*** | [0.05, 0.10] | 12757 |
| Others | 0.037 | [-0.02, 0.10] | 0.144*** | [0.11, 0.18] | 15938 |
| **Household size** |  |  |  |  |  |
| 1--4 |  |  |  |  | 13141 |
| 5--7 | -0.068** | [-0.11, -0.02] | -0.106*** | [-0.13, -0.08] | 11803 |
| 8+ | -0.129*** | [-0.18, -0.08] | -0.201*** | [-0.23, -0.17] | 10919 |
| **Place of residence** |  |  |  |  |  |
| Urban |  |  |  |  | 17135 |
| Rural | 0.038 | [-0.03, 0.10] | 0.031 | [-0.00, 0.06] | 9803 |
| **Wealth quintile/MPCE quintile** |  |  |  |  |  |
| Poorest |  |  |  |  | 4657 |
| Poorer | -0.022 | [-0.08, 0.03] | 0.120*** | [0.09, 0.15] | 7267 |
| Middle | -0.070* | [-0.13, -0.01] | 0.182*** | [0.15, 0.21] | 10330 |
| Richer | -0.118*** | [-0.19, -0.05] | 0.236*** | [0.20, 0.27] | 14868 |
| Richest | -0.307*** | [-0.39, -0.22] | 0.325*** | [0.28, 0.37] | 23424 |
| **Type of facility** |  |  |  |  |  |
| Private health facility |  |  |  |  | 31648 |
| Public health Facility | -1.577*** | [-1.64, -1.51] | -1.999*** | [-2.02, -1.97] | 2944 |
| **Year of Births** |  |  |  |  |  |
| 2014 |  |  |  |  | 14492 |
| 2015 | -0.023 | [-0.18, 0.13] | -0.023 | [-0.10, 0.05] | 12983 |
| 2016 | -0.021 | [-0.17, 0.13] | -0.022 | [-0.09, 0.05] | 12384 |
| 2017 | -0.018 | [-0.17, 0.13] | -0.01 | [-0.08, 0.06] | 12417 |
| 2018 | -0.028 | [-0.17, 0.12] | -0.03 | [-0.10, 0.04] | 11668 |
| 2019 | -0.043 | [-0.19, 0.10] | -0.021 | [-0.09, 0.05] | 11603 |
| 2020 | -0.158* | [-0.31, -0.00] | -0.080* | [-0.15, -0.01] | 10279 |
| 2021 | -0.166 | [-0.38, 0.05] | -0.042 | [-0.14, 0.06] | 10879 |
| Constant | 2.782*** | [2.61, 2.96] | 9.261*** | [9.18, 9.34] |  |

Note: \*\*\*p < 0.01, \*\*p < 0.05, \*p < 0.10 (indicates statistically significant)

We also estimated the predicted mean OOP payment for institutional delivery. The mean OOP payment was lower when compared to the predicted mean OOP payment in each category of socio-demographic and economic characteristics. For instance, the mean OOP payment was underestimated by 23% from the predicted mean OOP payment among women with higher education. For women belonging to the richest quintile, the mean OOP payment was underestimated by 24% from the predicted mean OOP payment. Likewise, in the case of private health centres, the mean OOP payment for institutional delivery was underestimated by 27% from the predicted mean OOP payment after controlling for socio-economic and demographic characteristics. The underestimation of mean OOP payment from the predicted mean OOP payment for institutional delivery did not show consistent pattern with time. Further, the deviation for the same is high by background characteristics and across all the states from NFHS-5 to NSS 75^th^ round. S4 Table shows the regression results of two-part model and the predicted OOP payment for NSS, 2018. In general, the pattern is similar but the estimates differ largely.

S5 Table presents the cases that had error in recording of responses on OOP payment. In NFHS survey, investigators were trained to report 9998 for those reported do not know to specific component of OOP questions. However, we found large number of data error in questions on OOP payment. Appendix 1 presents these cases with entries of 88, 998, 9998, 9888, 99999, 9999, 999, 99, that were supposed for missing and do-not know cases. Also, there were cases where a specific number was assigned for many/each of the component (99990).

## Discussion and Conclusion

Estimates of OOP payment on health care from population-based surveys are usually derived from data on total expenditure and reimbursement of households. The NFHS-4, 2015-16, for the first time, canvassed a set of questions on OOP payment of institutional delivery and it was also collected in NFHS-5,2019-21. Based on these questions, estimates of OOP payment were provided across the states and socio-economic groups. Data on OOP payment was collected for the last births to mothers during a period of five years preceding the survey. For example, if the survey in the state was conducted in December 2020, then the OOP payment for birth was recorded for 2016, 2017, 2018, 2019, and 2020 with a mid-period reference year of 2018. On the other hand. we have health surveys from NSS from which OOP estimates on institutional delivery is routinely derived. Both the surveys are large scale, undertaken in close time period and representative for states of India. In this context, we compare the estimates of OOP across these surveys. The following are the salient findings.

First, we noticed some error in recording the OOP data in NFHS-5 surveys. These records need to be edited before scientific analyses. These errors may yield bias estimates at national, state and district level. These errors are caused by investigators and not corrected in secondary edit. Conditional on those who had incurred OOP, the estimates of OOP were over estimated by 21% (1752 rupees) if these errors were not corrected. The smaller the geographical region (district), larger is the extent of error. In some states, these errors overestimate OOP by over 25%. Second, we found stark difference in the distribution of OOP payment from both the surveys. The simple density plot of OOP distribution from both the surveys revealed variations across surveys and higher skewness in the NFHS based distribution. Third, we observed a pattern in distribution of OOP payment based on economic conditions of the households. Taking NSS estimates as standard, the mean OOP of poorest quantile from NFHS were under-estimated while among richest quintile it was overestimated. The quantum of underestimation and overestimation was as high as 20%. We also found variations in OOP and non-overlapping confidence intervals across socio-demographic characteristics in both the surveys. The variation of NFHS-5 and NSS health survey was higher among less educated and poorer women. Third, our results suggest that the OOP estimates on institutional delivery varies largely across states of India. The extent of variations in OOP was as high as 129% in Arunachal Pradesh, 85% in the state of Manipur, and 82% in the state of Mizoram. Fourth, besides the aggregate OOP, the share of the components of OOP payment on institutional delivery derived from NFHS is not consistent with that derived from NSS. The analyses of OOP by its component such as medicine, hospital stay, tests, medicine and other costs are not consistent across surveys. Thus, considering the NSS estimates as the gold standard, the estimates of OOP payment on institutional delivery in NFHS-5 seem to yield inconsistent estimates for India and states of India.

Estimates of OOP payment derived from NFHS and NSS surveys are not consistent. The observable differences can be attributed to the fundamental differences in the structure of the questions canvassed in NSS and NFHS. The NSS asked the respondents two questions: expenditure on hospitalization and reimbursement, consequently from which the OOP payment was derived. In the case of NFHS-5, the questions on OOP were direct. It was expected that the respondents would do their own estimation and describe their OOP payment on institutional delivery. Given the level of educational qualification of the respondents and the likelihood of recall bias, it might not have been easy for the respondents to calculate the expenditure and provide accurate information on OOP payment. While some variations on OOP payment across surveys may be inevitable, however, high inconsistencies between the estimates of OOP for public health centres (catering to majority of deliveries) is a matter of concern. Besides, the sample size for NFHS is at least three times larger than the sample size of NSS. It is expected that the estimates derived from NFHS would be robust and consistent. But the distribution of OOP payments of institutional delivery for NFHS brings in light the fact that the large sample size is inadequate in providing with reliable estimates.

The following were the limitations of the study. First, we relied solely on OOP payment because there was no data on medical expenditure and reimbursement for expenses on institutional birth in NFHS-5. Secondly, we acknowledge that there may have been recall bias in reporting of the OOP payment, over the period of five years that could have affected the estimates despite the large sample size in NFHS-5. Third, estimates of catastrophic health expenditure on institutional birth could not be derived from the OOP payment due to the non-availability of consumption expenditure of households.

There are at least two implications of this study. First, a direct question on OOP is not easy for the respondents to provide an answer. The respondents are a heterogeneous group, with varying levels of educational attainment and belong to different cultural settings. Rather than asking direct questions from the respondents about their OOP payments it is better to ask them in a structured stepwise manner, which will leave the surveyors with a method to cross check the estimates, if they are correctly being specified or not. Hence the questions on OOP need to be modified and should follow a two-step procedure. In the first step, information on total expenditure may be collected, and in the second step, reimbursement data may be collected. It is recommended to use the NSS-based question on expenditure that is more standard and provides reliable estimates. Secondly, estimates of OOP payment should be price adjusted following data collection. Making price adjusted estimates is a post data collection exercise, like many other variables, such as wealth index, that are used in NFHS. Price-adjusted estimates are also recommended for evidence-based planning and research. It may be mentioned that since 2010, the Government of India has been publishing state-specific Consumer Price Index (CPI) by rural and urban areas on a monthly basis. Since data in NFHS was collected over a period of seven years, it is necessary to adjust the prices for reliable estimates.

## Data Availability

The data is freely available from https://dhsprogram.com/data/dataset/India_Standard-DHS_2020.cfm?flag=0 and http://microdata.gov.in/nada43/index.php/home

https://dhsprogram.com/data/dataset/India_Standard-DHS_2020.cfm?flag=0

## Appendix A. Supplementary data

## Declarations

### Ethics approval and consent to participate

The study used a secondary dataset which is freely available in the public domain. The survey agencies had obtained the prior consent from the respondents. The local ethics committee of the International Institute for Population Sciences ruled that no formal ethics approval was required to carry out research using this data source.

### Consent for publication

Not applicable.

## Acknowledgement

This paper was written as part of DataQi project of the Population Council funded by Bill & Melinda Gates Foundation (grant # OPP1194597). The views expressed herein are those of the authors and do not necessarily reflect the official policy or position of the Bill & Melinda Gates Foundation and/or Population Council. The authors would like to acknowledge the support from National Data Quality Forum (NDQF) in providing feedback to initial draft of the paper.

