## Supplementary File for "Data Quality of Out-Of-Pocket Payment on Institutional Delivery in India"

**S1(a) Table: Question asked on OOP on maternal care in NFHS-5, 2019-21**

| 448A | How much did it cost you out of your pocket for transportation? |
| --- | --- |
| 448B | How much did it cost you out of your pocket during delivery on: |
|  | a.hospital stay |
|  | b.tests done? |
|  | c.  medicines? |
|  | d. other costs? |
| 448C | CHECK 448B a-d: |
| 449 | How much in total did it cost you out of your pocket for this delivery? |
| 450 | CHECK 448A, 448B a-d, AND 449 |
| 451 | How was the out of pocket cost met? |
| 452 | Did you receive any financial assistance for delivery care? |
| 453 | From where did you get assistance? |
|  | a. Janani Suraksha Yojana |
|  | b. Other Government Scheme |
|  | x . Other |
| 454 | How many days after delivery did you receive the financial assistance under JSY? |
| 455 | What was the total amount that you Received? |
| 456 | How long after (NAME) was delivered did you stay in the health facility? |

**S1(b): Question asked on OOP on maternal care in NSS, 2018**

| **[6] particulars of medical treatment received as in-patient of a medical institution during the last 365 days** | | | | | | | | |
| --- | --- | --- | --- | --- | --- | --- | --- | --- |
| 1. | sr1. no. of the hospitalisation case | | | **1** | **2** | **3** | **4** | **5** |
| 2. | srl. no. of member (as in col. 1, block 4A/5 & 4B) hospitalised | | |  |  |  |  |  |
| 3. | age (years) (as in col.5, block 4A & 4B/ col.4, block 5) | | |  |  |  |  |  |
| 4. | nature of ailment (code list on pages 14-15)* | | |  |  |  |  |  |
| 5. | nature of treatment (code) | | |  |  |  |  |  |
| 6. | type of medical institution (code) | | |  |  |  |  |  |
| 7. | **if code is 2 or 3 in item 6**, reason for not availing govt./public  hospital | | |  |  |  |  |  |
| 8. | type of ward (free -1, paying general -2, paying special -3) | | |  |  |  |  |  |
| 9. | when admitted (code) | | |  |  |  |  |  |
| 10. | when discharged (code) | | |  |  |  |  |  |
| 11. | duration of stay in hospital (days) | | |  |  |  |  |  |
| **details of medical services received** (not received -1; received: free -2, partly free -3, on payment -4) | | | | | | | | |
| 12. | surgery | | |  |  |  |  |  |
| 13. | medicine | | |  |  |  |  |  |
| 14. | X-ray/ECG/EEG/Scan | | |  |  |  |  |  |
| 15. | other diagnostic tests | | |  |  |  |  |  |
| 16. | whether treated on medical advice before hospitalisation (yes -1,  no-2) | | |  |  |  |  |  |
| **if 1 in item 16** | | 17. | nature of treatment (code) |  |  |  |  |  |
|  |  | 18. | level of care (code) |  |  |  |  |  |
|  |  | 19. | duration of treatment (days) |  |  |  |  |  |
| 20. | whether treatment on medical advice continued after discharge  from hospital (yes -1, no-2) | | |  |  |  |  |  |
| **if 1 in item 20** | | 21. | nature of treatment (code) |  |  |  |  |  |
|  |  | 22. | level of care (code) |  |  |  |  |  |
|  |  | 23. | duration of treatment (days) |  |  |  |  |  |

| **S2 Table: Mean OOP payment on institutional delivery background characteristics in India, NFHS-5, 2019-21 & NSS, 2018** | | | | | | |  |  |
| --- | --- | --- | --- | --- | --- | --- | --- | --- |
|  | **NFHS-5, 2019-21** | | | | **NSS 75th round, 2018** | | | |
| **Background Characteristics** | **Public** | | **Private** | | **Public** | | **Private** | |
|  | **Mean** | **95%CI** | **Mean** | **95%CI** | **Mean** | **95%CI** | **Mean** | **95%CI** |
| **India** | **2894** | **[2843-2945]** | **23231** | **[22906-23557]** | **2738** | **[2644-2832]** | **23460** | **[22141-24779]** |
| **Education** |  |  |  |  |  |  |  |  |
| No education | 2249 | [2166,2332] | 16785 | [16093,17477] | 1964 | [1812,2117] | 18113 | [15927,20299] |
| Primary | 2602 | [2490,2713] | 19032 | [18057,20006] | 2294 | [2160,2429] | 19066 | [17571,20561] |
| Middle/secondary | 3301 | [3228,3374] | 22923 | [22493,23353] | 2856 | [2708,3003] | 20823 | [19634,22012] |
| Higher secondary & above | 4272 | [4065,4479] | 29612 | [29003,30222] | 4035 | [3719,4351] | 27434 | [24880,29988] |
| **Religion** |  |  |  |  |  |  |  |  |
| Hindu | 3038 | [2978,3098] | 24443 | [24074,24813] | 2652 | [2552,2751] | 23460 | [21806,25113] |
| Muslim | 3268 | [3126,3411] | 22608 | [21813,23402] | 2885 | [2659,3112] | 22412 | [20898,23926] |
| Others | 4032 | [3754,4310] | 28975 | [27616,30334] | 3944 | [3086,4802] | 26881 | [24305,29456] |
| **Caste** |  |  |  |  |  |  |  |  |
| SC/ST | 2741 | [2666,2816] | 21340 | [20760,21920] | 2363 | [2244,2482] | 20653 | [19297,22010] |
| OBC | 3067 | [2987,3148] | 24699 | [24204,25195] | 2706 | [2578,2834] | 22813 | [21786,23840] |
| Others | 3811 | [3671,3951] | 26006 | [25380,26631] | 3530 | [3226,3835] | 25716 | [22261,29171] |
| **Household size** |  |  |  |  |  |  |  |  |
| 1--4 | 3473 | [3366,3581] | 26317 | [25659,26975] | 2744 | [2579,2908] | 24190 | [22760,25620] |
| 5--7 | 3003 | [2935,3071] | 24444 | [23969,24919] | 2752 | [2618,2887] | 23658 | [21069,26248] |
| 8+ | 2887 | [2784,2991] | 22035 | [21463,22606] | 2699 | [2507,2892] | 22195 | [20907,23483] |
| **Place of residence** |  |  |  |  |  |  |  |  |
| Urban | 3561 | [3419,3704] | 26722 | [26116,27327] | 3753 | [3462,4045] | 27048 | [24176,29920] |
| Rural | 2969 | [2913,3025] | 22693 | [22349,23037] | 2500 | [2408,2592] | 20932 | [20044,21819] |
| **Wealth quintile/MPCE quintile** |  |  |  |  |  |  |  |  |
| Poorest | 2371 | [2296,2446] | 17050 | [16295,17805] | 2216 | [2089,2342] | 18716 | [17464,19968] |
| Poorer | 2985 | [2894,3077] | 19543 | [18885,20200] | 2668 | [2491,2844] | 20091 | [18938,21243] |
| Middle | 3293 | [3184,3401] | 21814 | [21224,22404] | 2754 | [2585,2923] | 24595 | [18419,30771] |
| Richer | 3587 | [3446,3728] | 24536 | [23945,25127] | 2975 | [2766,3184] | 24542 | [22893,26192] |
| Richest | 3935 | [3718,4153] | 28832 | [28180,29484] | 3647 | [3209,4085] | 26296 | [24925,27666] |
| **Caesarean Delivery** |  |  |  |  |  |  |  |  |
| Yes | 6746 | [6522,6970] | 32903 | [32397,33410] | 6467 | [6017,6916] | 31803 | [30840,32766] |
| No | 2409 | [2366,2451] | 16024 | [15628,16420] | 2232 | [2153,2311] | 15669 | [13239,18098] |

| **S3 Table: Mean estimates of OOP payment on institutional delivery (₹) by major states of India** | | | | | | | | |
| --- | --- | --- | --- | --- | --- | --- | --- | --- |
|  | **NFHS-5, 2019-21** | | | | **NSS 75th round, 2018** | | | |
|  | **Public** | | **Private** | | **Public** | | **Private** | |
| **States** | **Mean** | **95%CI** | **Mean** | **95%CI** | **Mean** | **95%CI** | **Mean** | **95%CI** |
| **India** | **2894** | **[2843-2945]** | **23231** | **[22906-23557]** | **2738** | **[2644-2832]** | **23460** | **[22141-24779]** |
| Jammu & Kashmir | 5714 | [5397,6030] | 25579 | [22610,28548] | 5153 | [4594,5712] | 28493 | [24323,32663] |
| Himachal Pradesh | 4066 | [2907,5226] | 30232 | [26481,33983] | 4075 | [3546,4604] | 27234 | [21275,33194] |
| Punjab | 4142 | [3768,4516] | 27223 | [25752,28694] | 4216 | [3087,5345] | 21093 | [19075,23110] |
| Chandigarh | 5569 | [3322,7817] | 46752 | [25149,68356] | 5446 | [3180,7712] | 44661 | [37164,52158] |
| Uttarakhand | 3474 | [2916,4032] | 28645 | [25788,31503] | 2816 | [2277,3355] | 23448 | [19111,27786] |
| Haryana | 1727 | [1534,1921] | 23031 | [21941,24120] | 2068 | [1709,2428] | 34598 | [7723,61473] |
| Delhi | 2708 | [2212,3204] | 33975 | [31558,36391] | 4144 | [2417,5871] | 33492 | [27548,39437] |
| Rajasthan | 2190 | [2055,2324] | 17531 | [16579,18482] | 2441 | [2152,2730] | 16095 | [13719,18470] |
| Uttar Pradesh | 2445 | [2321,2569] | 20800 | [20194,21406] | 1827 | [1603,2051] | 21326 | [19136,23515] |
| Bihar | 2864 | [2713,3016] | 19795 | [18830,20759] | 2554 | [2317,2792] | 15986 | [13989,17983] |
| Sikkim | 9099 | [7454,10744] | 21714 | [14633,28796] | 6108 | [4554,7662] | 19866 | [14871,24861] |
| Arunachal Pradesh | 11315 | [10179,12451] | 39222 | [32138,46307] | 3949 | [3093,4806] | 13235 | [7185,19286] |
| Nagaland | 6103 | [5316,6889] | 21099 | [18423,23774] | 4334 | [2978,5690] | 16188 | [11775,20602] |
| Manipur | 15286 | [14400,16171] | 37477 | [34742,40212] | 9034 | [8167,9900] | 31634 | [27140,36128] |
| Mizoram | 7234 | [5859,8608] | 23753 | [19113,28392] | 2399 | [2021,2777] | 5825 | [2834,8815] |
| Tripura | 7217 | [6591,7842] | 27026 | [24226,29826] | 6306 | [5358,7254] | 33105 | [29374,36836] |
| Meghalaya | 3561 | [3089,4032] | 29801 | [24826,34776] | 2778 | [2199,3357] | 12369 | [8259,16479] |
| Assam | 5945 | [5653,6236] | 33484 | [31231,35737] | 3732 | [3312,4152] | 28589 | [23846,33331] |
| West Bengal | 2881 | [2632,3131] | 22325 | [21130,23519] | 2979 | [2507,3452] | 24813 | [22824,26801] |
| Jharkhand | 2254 | [2078,2431] | 22587 | [20772,24401] | 2484 | [1981,2987] | 16710 | [13919,19501] |
| Orissa | 4635 | [4376,4894] | 25553 | [24132,26974] | 3789 | [3479,4098] | 23109 | [20902,25315] |
| Chhattisgarh | 2114 | [1909,2318] | 24235 | [22203,26266] | 1834 | [1442,2226] | 20604 | [18221,22987] |
| Madhya Pradesh | 1798 | [1664,1933] | 28408 | [26629,30186] | 1584 | [1353,1815] | 22241 | [18184,26298] |
| Gujarat | 1781 | [1520,2042] | 17286 | [16287,18285] | 1951 | [1597,2306] | 16440 | [14701,18180] |
| Daman & Diu | 693 | [519,867] | 20437 | [17396,23478] | 1374 | [983,1766] | 28943 | [16459,41426] |
| Dadra & Nagar Haveli |  |  |  |  | 838 | [380,1295] | 14243 | [8298,20189] |
| Maharashtra | 3252 | [3010,3495] | 25177 | [23741,26614] | 3168 | [2768,3568] | 22880 | [20928,24832] |
| Andhra Pradesh | 3005 | [2605,3405] | 23632 | [22321,24943] | 2853 | [2333,3374] | 24227 | [22346,26109] |
| Karnataka | 5108 | [4741,5474] | 26803 | [25317,28290] | 4044 | [3598,4489] | 23036 | [21051,25021] |
| Goa | 4104 | [3453,4754] | 38187 | [33287,43086] | 4446 | [3404,5487] | 36390 | [29056,43725] |
| Lakshadweep | 2631 | [1412,3850] | 51421 | [41879,60964] | 5696 | [3693,7698] | 61331 | [48559,74103] |
| Kerala | 6991 | [6261,7721] | 35620 | [34066,37175] | 8642 | [7287,9998] | 30766 | [28983,32549] |
| Tamil Nadu | 3562 | [3350,3774] | 37421 | [35739,39103] | 3643 | [3316,3970] | 34000 | [30448,37552] |
| Pondicherry | 3456 | [2731,4181] | 24133 | [21013,27252] | 3016 | [2199,3832] | 38178 | [31844,44511] |
| Andaman & Nicobar Islands | 3106 | [2104,4108] | 50383 | [39903,60862] | 2060 | [1242,2879] | 53830 | [43126,64535] |
| Telangana | 4204 | [3871,4538] | 25086 | [23841,26331] | 3305 | [2709,3901] | 31854 | [29235,34474] |
| Ladakh | 4263 | [3711,4814] | 14276 | [-2951,31504] |  |  |  |  |
| * Daman & Diu and Dadar & Nagar haveli are combined in NFHS-5 raw data, hence the estimates are combined | | | | | | | | |

| **S4 Table: Results of the two-part regression model and predicted OOPE on institutional delivery in India, NSS (2018)** | | | | | |
| --- | --- | --- | --- | --- | --- |
| **Background characteristics** | **β (logit)** | **95% CI** | **β (OLS)** | **95% CI** | **Mean OOP payment** |
| **Education** |  |  |  |  |  |
| No education ® |  |  |  |  | 8935 |
| Primary | 0.19 | [-0.11,0.49] | -0.023* | [-0.05,-0.00] | 8750 |
| Middle/secondary | -0.025 | [-0.29,0.24] | 0.112*** | [0.09,0.13] | 13435 |
| Higher secondary & above | -0.184 | [-0.46,0.10] | 0.212*** | [0.19,0.24] | 25007 |
| **Religion** |  |  |  |  |  |
| Hindu ® |  |  |  |  | 14349 |
| Muslim | 0.295 | [-0.03,0.62] | 0.011 | [-0.01,0.03] | 14768 |
| Others | -0.305* | [-0.56,-0.04] | 0.162*** | [0.13,0.19] | 21430 |
| **Caste** |  |  |  |  |  |
| SC/ST ® |  |  |  |  | 9002 |
| OBC | 0.533*** | [0.28,0.78] | 0.089*** | [0.07,0.11] | 15133 |
| Others | 0.129 | [-0.11,0.37] | 0.170*** | [0.15,0.19] | 21275 |
| **Household size** |  |  |  |  |  |
| 1--4 ® |  |  |  |  | 13927 |
| 5--7 | -0.034 | [-0.24,0.17] | 0.088*** | [0.07,0.11] | 14943 |
| 8+ | -0.219 | [-0.52,0.08] | 0.100*** | [0.07,0.13] | 15299 |
| **Place of residence** |  |  |  |  |  |
| Urban ® |  |  |  |  | 12459 |
| Rural | -0.661*** | [-0.87,-0.46] | 0.027** | [0.01,0.04] | 21256 |
| **Wealth quintile/MPCE quintile** |  |  |  |  |  |
| Poorest ® |  |  |  |  | 9248 |
| Poorer | 0.084 | [-0.25,0.42] | 0.116*** | [0.09,0.14] | 11719 |
| Middle | 0.002 | [-0.34,0.34] | 0.154*** | [0.13,0.18] | 14162 |
| Richer | -0.131 | [-0.46,0.20] | 0.199*** | [0.17,0.23] | 17391 |
| Richest | -0.604*** | [-0.92,-0.29] | 0.280*** | [0.25,0.31] | 23642 |
| **Type of facility** |  |  |  |  |  |
| Private health facility ® |  |  |  | [0.00,0.00] | 5073 |
| Public health Facility | 0.739*** | [0.53,0.95] | 1.855*** | [1.84,1.87] | 37478 |
| Constant | 5.380*** | [5.02,5.74] | 7.401*** | [7.37,7.43] |  |
| Note: ***p < 0.01, **p < 0.05, *p < 0.10 (indicates statistically significant) | | | | |  |

| **S5 Table: Illustration of error in OOP Data, NFHS-5** | | | |  |  |  |  |
| --- | --- | --- | --- | --- | --- | --- | --- |
| Sr no | Case ID | Transportation | Hospital stay | Test done | Medicine | Other cost | Total |
| 1 | 0919000364 02 | 98 | 98 | 98 | 98 | 99998 |  |
| 2 | 0919000358 07 | 98 | 98 | 98 | 98 | 6000 |  |
| 3 | 1023001513 02 | 98 | did not pay | did not pay | did not pay | did not pay | 1200 |
| 4 | 2749903863 03 | 98 | 98 | 98 | 98 | 30000 |  |
| 5 | 2749902531 02 | 98 | 5000 | 10000 | 98 | 12000 |  |
| 6 | 0812000145 04 | 99 | 9 | 9 | 9 | 9 |  |
| 7 | 0607503311 02 | 9998 | 99998 | 99998 | 9998 | 99998 |  |
| 8 | 0607601951 04 | 9998 | 9998 | 9998 | 9998 | 9998 |  |
| 9 | 0607702038 02 | 9998 | 9998 | 9998 | 99998 | 25000 |  |
| 10 | 0608301590 02 | 9998 | 99998 | 99998 | 9998 | 6000 |  |
| 11 | 0783900288 04 | 9998 | did not pay | did not pay | did not pay | 9998 |  |
| 12 | 0914700125 01 | 9998 | did not pay | did not pay | did not pay | 9998 |  |
| 13 | 0915004530 02 | 9998 | did not pay | did not pay | 9998 | 3500 |  |
| 14 | 0919801396 04 | 9998 | 9998 | 99998 | 99998 | 99998 |  |
| 15 | 1021604301 04 | 9998 | 2000 | 1500 | 2000 | did not pay |  |
| 16 | 1023801801 04 | 9998 | 99998 | 99998 | 99998 | 500 |  |
| 17 | 1224500798 02 | 9998 | 9998 | 9998 | 9998 | 9998 |  |
| 18 | 1224602094 02 | 9998 | 99998 | 4000 | 3000 | 99998 |  |
| 19 | 1225200185 02 | 9998 | 9998 | 99998 | 99998 | 99998 |  |
| 20 | 1280201396 02 | 9998 | 9998 | 9998 | 9998 | 9998 |  |
| 21 | 1280901475 02 | 9998 | 99998 | 99998 | 99998 | 99998 | 99998 |
| 22 | 2036302339 02 | 9998 | did not pay | did not pay | did not pay | did not pay | did not pay |
| 23 | 2036900353 03 | 9998 | did not pay | did not pay | did not pay | did not pay | 5000 |
| 24 | 2345501783 04 | 9998 | 99998 | 99998 | 99998 | 99998 | 36000 |
| 25 | 3463601414 05 | 9998 | 99998 | 99998 | 99998 | 99998 | 25000 |
| 26 | 3688802820 02 | 9998 | did not pay | did not pay | did not pay | did not pay | 1000 |
| 27 | 2035703280 01 | 9999 | 9999 | 800 | 9999 | 200 |  |
| 28 | 2035700790 04 | 9999 | 9999 | 9999 | 9999 | 500 |  |
| 29 | 2035700708 03 | 9999 | 9999 | 9999 | 9999 | 500 |  |
| 30 | 2035702313 04 | 9999 | 9999 | 9999 | 9999 | 500 |  |
| 31 | 2750100945 04 | 9999 | 35000 | 1000 | 2000 | did not pay |  |
| 32 | 3158701704 02 | 9999 | 99998 | 99998 | 99998 | 99998 | 99998 |
| 33 | 0505702002 03 | 99990 | 99990 | 99990 | 99990 | 99990 |  |
| 34 | 0810802180 05 | 99990 | did not pay | did not pay | did not pay | did not pay | did not pay |
| 35 | 0812601127 04 | 99990 | 99990 | 99990 | 99990 | did not pay |  |
| 36 | 0913202577 02 | 99990 | 99990 | 600 | 800 | 1000 |  |
| 37 | 0914500240 04 | 99990 | 99998 | 99998 | 99998 | 99998 | 25000 |
| 38 | 1022500532 05 | 99990 | 99990 | 99990 | 99990 | 500 |  |
| 39 | 1022702632 05 | 99990 | 99990 | 99990 | 200 | 1000 |  |
| 40 | 1022702632 09 | 99990 | 99990 | 99990 | 99990 | 1000 |  |
| 41 | 1023802419 01 | 99990 | 99990 | 99990 | 500 | 1500 |  |
| 42 | 1023803897 02 | 99990 | 99990 | 500 | 99990 | 2000 |  |
| 43 | 1023803893 04 | 99990 | 99990 | 500 | 1000 | 4000 |  |
| 44 | 1124300446 02 | 99990 | 99990 | 600 | 500 | 10000 |  |
| 45 | 1124300442 03 | 99990 | 99990 | 4000 | 3000 | 50000 |  |
| 46 | 1528303612 02 | 99990 | 1500 | 99998 | 2000 | 1000 |  |
| 47 | 1729703055 03 | 99990 | did not pay | did not pay | did not pay | did not pay | did not pay |
| 48 | 1934301106 04 | 99990 | did not pay | did not pay | 2000 | 100 |  |
| 49 | 1934503602 04 | 99990 | did not pay | did not pay | 99990 | did not pay |  |
| 50 | 2956400716 06 | 99990 | 99990 | 1500 | 99990 | 99990 |  |
| 51 |  |  |  |  |  |  |  |
| 52 | 0992600744 02 | 150 | 98 | 98 | 98 | 5000 |  |
| 53 | 2749800661 02 | 100 | 98 | 98 | 98 | 1200 |  |
| 54 | 2749903898 04 |  | 98 | 98 | 2000 | 17000 |  |
| 55 | 2749902528 02 | 100 | 98 | 98 | 98 | 40000 |  |
| 56 | 2855202342 05 | 200 | 98 | 98 | 98 | 5000 |  |
| 57 | 0916200973 01 | did not pay | 99 | did not pay | did not pay | 200 |  |
| 58 | 0202402997 02 | 99998 | 9998 | 9998 | 9998 | 9998 |  |
| 59 | 0607701948 02 | did not pay | 9998 | 9998 | 9998 | 15000 |  |
| 60 | 0810704555 02 | 800 | 9998 | 9998 | 2500 | 10000 |  |
| 61 | 0913700210 04 | 55000 | 9998 | 3000 | 9998 | 9998 |  |
| 62 | 0913700260 02 | 200 | 9998 | 9998 | 9998 | 9998 |  |
| 63 | 0913700202 02 | 25000 | 9998 | 2500 | 2000 | 9998 |  |
| 64 | 0914601124 04 | 500 | 9998 | 9998 | 9998 | 9998 |  |
| 65 | 0992803545 08 | 300 | 9998 | 9998 | 500 | 9998 |  |
| 66 | 1023601157 04 | 99998 | 9998 | 99998 | 99998 | 50000 |  |
| 67 | 1023601153 01 | 350 | 9998 | 99998 | 9998 | 550 |  |
| 68 | 1729800722 02 | 99998 | 9998 | 9998 | 99998 | 99998 |  |
| 69 | 2036302331 01 | 2500 | 9998 | 99998 | 99998 | 30000 |  |
| 70 | 2036303646 10 | 1000 | 9998 | 1500 | 9998 | did not pay |  |
| 71 | 2036303692 04 | 200 | 9998 | did not pay | 9998 | 45000 |  |
| 72 | 2240401954 02 | 100 | 9998 | 300 | 2000 | 500 |  |
| 73 | 2751200698 02 | 300 | 9998 | 4000 | 99998 | 99998 |  |
| 74 | 2786904122 02 | did not pay | 9998 | 99998 | 99998 | 99998 |  |
| 75 | 0914403962 02 | 1000 | 9999 | 99998 | 2000 | did not pay |  |
| 76 | 0919104335 04 | 500 | 9999 | 99998 | 99998 | 35000 |  |
| 77 | 1528203762 05 | 300 | 9999 | 99998 | 99998 | 2000 |  |
| 78 | 2035700692 04 | 500 | 9999 | 9999 | 9999 | 500 |  |
| 79 | 2035700739 02 | 1000 | 9999 | 9999 | 9999 | 200 |  |
| 80 | 2035700777 04 | 500 | 9999 | 9999 | 9999 | 500 |  |
| 81 | 1832500987 02 | 2000 | 98000 | 98000 | 98000 | 29000 |  |
| 82 | 0202803285 04 | 1000 | 99990 | 30000 | 10000 | 20000 |  |
| 83 | 0505702670 02 | 200 | 99990 | 3000 | 1000 | 99990 |  |
| 84 | 0506200187 07 | 200 | 99990 | 4000 | 5000 | 99998 |  |
| 85 | 0784001776 02 | 1000 | 99990 | 500 | 800 | did not pay |  |
| 86 | 0811100731 11 | 30 | 99990 | 99990 | 99990 | did not pay |  |
| 87 | 0811100123 04 | 200 | 99990 | 1400 | did not pay | did not pay |  |
| 88 | 1022302899 07 | 99998 | 99990 | 99990 | 99990 | 99990 |  |
| 89 | 1022504367 02 | 300 | 99990 | 99990 | 99990 | 99990 |  |
| 90 | 1022500507 02 | 200 | 99990 | 99990 | 99990 | 1000 |  |
| 91 | 1022504170 02 | 500 | 99990 | 99990 | 99990 | 1000 |  |
| 92 | 1022700185 03 | 100 | 99990 | 99990 | 99990 | 500 |  |
| 93 | 1023701728 06 | 500 | 99990 | 1000 | 500 | 3000 |  |
| 94 | 1023803748 03 | 500 | 99990 | 500 | 500 | 3000 |  |
| 95 | 1023803803 04 | 300 | 99990 | 5000 | 4000 | 20000 |  |
| 96 | 1280903485 02 | 99998 | 99990 | 200 | 99990 | 99998 |  |
| 97 | 1729702738 02 |  | 99990 | 40000 | 20000 | 10000 |  |
| 98 | 1729702376 02 | 2200 | 99990 | 1500 | 36000 | 4000 |  |
| 99 | 1933804230 03 | 1 | 99990 | 99990 | 99998 | 99990 |  |
| 100 | 2138200159 02 | 5000 | 99990 | 70000 | 20000 | did not pay |  |
| 101 | 2854500947 02 | 300 | 99990 | 5000 | 7000 | 15000 |  |
| 102 | 2854502160 02 | did not pay | 99990 | 6000 | did not pay | 3000 |  |
| 103 | 2956402504 02 | 1000 | 99990 | 99990 | 99990 | 99990 |  |
| 104 | 2956402540 10 | 200 | 99990 | 1500 | 2000 | 1000 |  |
| 105 | 2956402578 04 | 40 | 99990 | 99990 | 99990 | 3000 |  |
| 106 | 2956503173 02 | 500 | 99990 | 99990 | 99990 | 99990 |  |
| 107 | 2956503180 04 | 100 | 99990 | 99990 | 99990 | 500 |  |
| 108 | 2956503131 02 |  | 99990 | 99990 | 99990 | 1000 |  |
| 109 | 3259900101 03 | 99998 | 99990 | 30000 | 30000 | 40000 |  |
| 110 | 2749801395 04 | 200 | 25000 | 98 | 98 | 98 |  |
| 111 | 2749801395 06 | 500 | 27000 | 98 | 98 | 98 |  |
| 112 | 2749800429 04 | 1000 | 35000 | 98 | 10000 | 98 |  |
| 113 | 0607503031 02 | 99998 | 99998 | 9998 | 9998 | 99998 |  |
| 114 | 0405503421 02 | 200 | 99998 | 9999 | 99998 | 5000 |  |
| 115 | 2036401792 11 | 1000 | 500 | 9999 | 9999 | 1000 |  |
| 116 | 1832502196 03 | did not pay | did not pay | 98000 | 98000 | 6000 |  |
| 117 | 1832502176 04 | did not pay | did not pay | 98000 | 98000 | 13000 |  |
| 118 | 0915900931 04 | 99998 | 99998 | 98888 | 60000 | did not pay |  |
| 119 | 0913203793 04 | 500 | 1 | 99990 | 99990 | did not pay |  |
| 120 | 1326504151 04 | did not pay | 99998 | 99990 | 99990 | 99998 |  |
| 121 | 1831603468 05 | 200 | 5000 | did not pay | 98 | 99998 |  |
| 122 | 0917800745 02 | 200 | did not pay | did not pay | 99 | 1000 |  |
| 123 | 0607701130 04 | 1000 | 1500 | 99998 | 9998 | 25000 |  |
| 124 | 0914603161 04 | 1500 | did not pay | did not pay | 9998 | 2000 |  |
| 125 | 1023601107 02 | 200 | did not pay | did not pay | 9998 | 700 |  |
| 126 | 1225500822 02 | did not pay | did not pay | did not pay | 9998 | 2000 |  |
| 127 | 1831802147 07 | 99998 | 99998 | 99998 | 9998 | 99998 |  |
| 128 | 1881103426 03 | 700 | 2000 | 500 | 9998 | 9998 |  |
| 129 | 1881800716 03 | did not pay | did not pay | did not pay | 9998 | 10000 |  |
| 130 | 2137000251 02 | did not pay | did not pay | 99998 | 9998 | 99998 |  |
| 131 | 2137702731 07 | did not pay | did not pay | did not pay | 9998 | 9998 |  |
| 132 | 2138500885 04 | did not pay | 99998 | 99998 | 9998 | 9998 |  |
| 133 | 1832502106 02 | did not pay | did not pay | did not pay | 98000 | 5000 |  |
| 134 | 0914801186 02 | did not pay | did not pay | did not pay | 1400 | 98 |  |
| 135 | 1528404104 01 | did not pay | did not pay | 400 | did not pay | 98 |  |
| 136 | 1831600460 01 | 300 | did not pay | did not pay | 900 | 98 |  |
| 137 | 1831603475 02 | did not pay | did not pay | did not pay | 3000 | 98 |  |
| 138 | 1831600101 05 | did not pay | 5000 | did not pay | 5000 | 98 |  |
| 139 | 1933104072 03 | did not pay | did not pay | did not pay | 500 | 98 |  |
| 140 | 2485902510 02 | 300 | 99998 | 99998 | 99998 | 99 |  |
| 141 | 0914604104 07 | 60000 | 20000 | 20000 | 20000 | 9998 |  |
| 142 | 0915904460 04 | 100 | 2000 | 2000 | 1000 | 9998 |  |
| 143 | 0917502138 02 | 500 | 3600 | 99998 | 99998 | 9998 |  |
| 144 | 1020304207 04 | 3500 | 1000 | 99998 | 99998 | 9998 |  |
| 145 | 1021701564 04 | 200 | did not pay | did not pay | 400 | 9998 |  |
| 146 | 1023702169 03 | 99998 | 99998 | did not pay | did not pay | 9998 |  |
| 147 | 1224501793 02 | 99998 | 99998 | 99998 | 99998 | 9998 |  |
| 148 | 1691600724 03 | 500 | did not pay | 1000 | 500 | 9998 |  |
| 149 | 1691802926 02 | 99998 | did not pay | 2000 | 2000 | 9998 |  |
| 150 | 1933903750 02 | 99998 | 99998 | 99998 | 99998 | 9998 |  |
| 151 | 2344300857 02 | did not pay | 500 | did not pay | did not pay | 9998 |  |
| 152 | 2344300890 02 | did not pay | 99998 | 99998 | 99998 | 9998 |  |
| 153 | 0914504175 04 | 99998 | 99998 | 99998 | 99998 | 98888 |  |
| 154 | 0783801464 08 | did not pay | did not pay | did not pay | did not pay | 99990 |  |
| 155 | 0784402239 03 | did not pay | did not pay | did not pay | did not pay | 99990 |  |
| 156 | 0813104413 06 | did not pay | 4000 | 99998 | 99998 | 99990 |  |
| 157 | 1022302877 02 | 100 | did not pay | did not pay | 99998 | 99990 |  |
| 158 | 2283001797 04 | 1000 | 3000 | 2000 | 5000 | 99990 |  |
| 159 | 2854500753 04 | 300 | did not pay | did not pay | did not pay | 99990 |  |
| 160 | 3362100423 04 | 10000 | 20000 | 5000 | 500 | 99990 |  |
| 161 | 0505602087 02 | did not pay | did not pay | did not pay | did not pay | 99000 |  |
| 162 | 0608301316 04 | did not pay | 99998 | 99998 | 99998 | 99000 |  |
| 163 | 0608300910 02 | 99998 | 99998 | 99998 | 99998 | 99000 |  |
| 164 | 0783802241 03 | 1000 | 20000 | 99998 | 99998 | 99000 |  |
| 165 | 3362700494 03 | did not pay | did not pay | did not pay | did not pay | 99000 |  |
| 166 | 0607001554 04 | 99998 | 99998 | 99998 | 99998 | 99998 | 99995 |
| 167 | 0784003509 02 | 99998 | 99998 | 99998 | 99998 | 99998 | 99995 |
| 168 | 0784101319 04 | did not pay | 99998 | 99998 | 99998 | 99998 | 99995 |
| 169 | 0784701316 03 | 250 | 99998 | 99998 | 99998 | 99998 | 99995 |
| 170 | 0784701475 10 | 99998 | 99998 | 99998 | 99998 | 99998 | 99995 |
| 171 | 0784701817 01 | 99998 | 99998 | 99998 | 99998 | 99998 | 99995 |
| 172 | 0809900270 02 | 1700 | 99998 | 99998 | 99998 | 99998 | 99995 |
| 173 | 0810002920 02 | 99998 | 99998 | 99998 | 99998 | 99998 | 99995 |
| 174 | 0813104418 04 | 50 | 99998 | 99998 | 99998 | 99998 | 99995 |
| 175 | 0914402314 03 | 99998 | 99998 | 99998 | 99998 | 99998 | 99995 |
| 176 | 0915503154 06 | did not pay | did not pay | did not pay | did not pay | did not pay | 99995 |
| 177 | 1224803145 02 | 99998 | 99998 | 99998 | 99998 | 99998 | 99995 |
| 178 | 2035702895 04 | 2200 | did not pay | did not pay | did not pay | did not pay | 99995 |
| 179 | 3360200185 04 | 50 | 99998 | 99998 | 99998 | 99998 | 99995 |
| 180 | 3362004030 02 | 100 | 99998 | 99998 | 99998 | 99998 | 99995 |
| 181 | 3362002369 03 | 99998 | 99998 | 99998 | 99998 | 99998 | 99995 |
| 182 | 3362400804 02 |  | did not pay | did not pay | did not pay | did not pay | 99995 |
| 183 | 3362603826 04 | 99998 | did not pay | did not pay | did not pay | did not pay | 99995 |
| 184 | 0202802043 06 | 99998 | 99998 | 99998 | 99998 | 99998 | 99990 |
| 185 | 3158703311 02 | 600 | 99998 | 99998 | 99998 | 99998 | 99990 |
| 186 | 3260102998 03 | did not pay | 99998 | 99998 | 99998 | 99998 | 99990 |
| 187 | 3689101827 03 | 50 | 99998 | 99998 | 99998 | 99998 | 99990 |
| 188 | 3689103647 04 | 100 | 99998 | 99998 | 99998 | 99998 | 99990 |
| 189 | 1021602339 03 | 99998 | 99998 | 99998 | 99998 | 99998 | 99988 |
| **Don't Know=9998, did not pay =0 | | | | | |  |  |

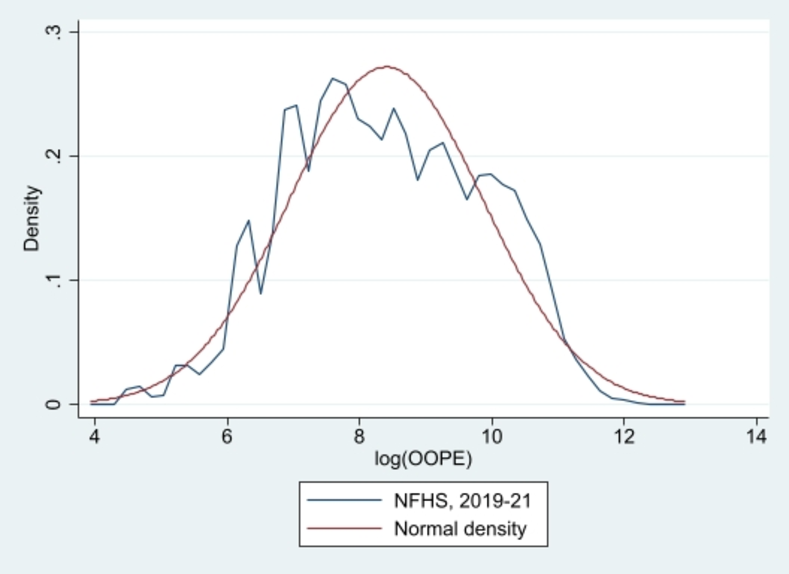

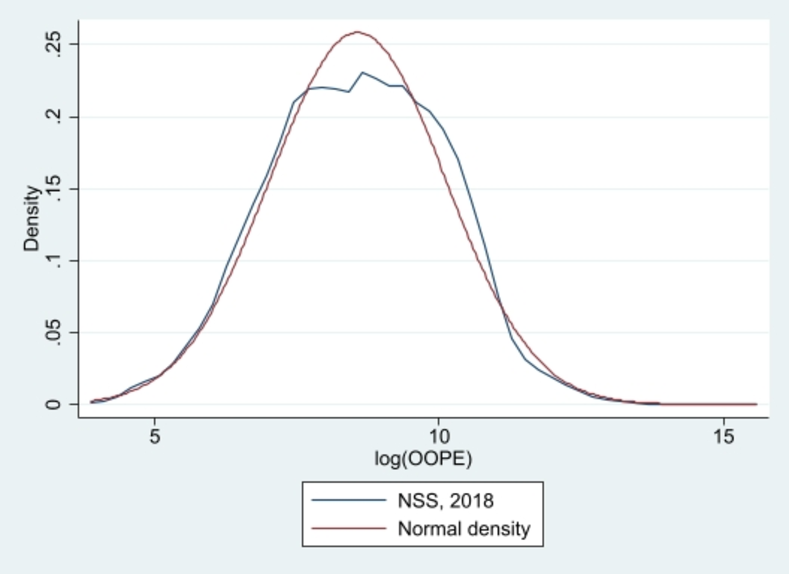

**S2 Fig: Density plot of log transformed OOP payment for NFHS, 2019-21 with Normal density**

**S1 Fig: Density plot of log transformed OOP payment of NSS, 2018 with Normal density**
